# A Qualitative Interview Study of General Practitioners’ Experiences of Managing Post-COVID-19 Syndrome

**DOI:** 10.1101/2024.04.23.24306074

**Authors:** Josefine Schulze, Lennart Lind, Alina Rojas, Laura Lüdtke, Jens Hensen, Corinna Bergelt, Martin Härter, Nadine Janis Pohontsch

## Abstract

**Background:** The management of the long-term sequelae of COVID-19 infection, known as post-COVID-19 syndrome (PCS), continues to challenge the medical community, largely due to a significant gap in the understanding of its aetiology, diagnosis and effective treatment.

**Aim:** To examine general practitioners’ (GPs) experiences of caring for patients with PCS and to identify unmet care needs and opportunities for improvement.

**Design and setting:** This study follows a qualitative design, using in-depth semi-structured telephone interviews with GPs (*N*=31) from across Germany.

**Method:** Interviews were audio-recorded, transcribed verbatim and analysed using qualitative content analysis.

**Results:** Patients with persistent symptoms after SARS-CoV-2 infection often consult their GPs as the first point of contact, with symptoms typically resolving within weeks. While ongoing symptomatic COVID-19 is perceived to be more common, the relevance of PCS to GP practices is considerable given its severe impact on patients’ functioning, social participation, and the substantial time required for patient care. GPs coordinate diagnosis and treatment, but face difficulties because of the unclear definition of PCS and difficulties in attributing symptoms, resulting in a cautious approach to ICD-10 coding. Interviewees highlight lengthy diagnostic pathways and barriers to accessing specialist care.

**Conclusion:** The findings confirm the high functional limitations and psychosocial burden of PCS on patients and the central role of GPs in their care. The study suggests a need for further research and health policy measures to support GPs in navigating diagnostic uncertainty, interprofessional communication and the limited evidence on effective treatments.

**How this fits in:** Post-COVID-19 syndrome has garnered attention in research and healthcare, but limited evidence on its causes and effective treatment challenges clinicians. This study illustrates the symptom-driven approaches to diagnosis and treatment adopted by general practitioners and their concerns about referring patients to specialist clinics. Greater collaboration and communication across sectors and disciplines is needed to meet the identified need for interprofessional care. Research should also focus on developing comprehensive differential diagnostic protocols, and health policy should address barriers to accessing specific outpatient services.

## Background

On 5 May 2023, the World Health Organisation declared the end of the COVID-19 pandemic as a global health emergency^1^. However, the virus remains active and a subset of patients continues to experience health problems long after their initial infection. Persistent symptoms following SARS-CoV-2 infection, known as long COVID, pose a challenge to the medical community. There are two phases of long COVID: the first is ongoing symptomatic COVID-19, which extends beyond the typical four-week duration after initial infection but resolves before the second phase, known as post-COVID-19 syndrome (PCS), where symptoms persist more than twelve weeks after infection^2,3^. While public interest was initially propelled by patient advocacy, the issue is now coming into the focus of international research and medical care^4^. Still, there is a significant gap in our understanding of treatment and parameters to support the diagnosis of PCS^5,6^.

In the German healthcare system, as in many other countries, general practitioners (GPs) are the first point of contact for patients with most health problems and serve as coordinators of their care. Nevertheless, little research is available on GPs’ experiences of caring for patients with PCS^7–9^. This study therefore examined GPs’ perceptions of the symptoms of PCS, their diagnostic approaches, collaboration with other disciplines and their assessment of the support, information, rehabilitation and treatment services available for these patients. By analysing GPs’ perspectives, we aimed to identify unmet health care needs and potential areas for improvement in the care of patients with PCS.

## Methods

We conducted semi-structured telephone interviews with GPs across Germany and utilised a qualitative content analysis approach to examine their experiential knowledge of managing patients with PCS^10^. Prior to data collection, the project was registered with the German Clinical Trials Registry (DRKS00029363).

### Recruitment

We employed a multi-faceted approach to recruit GPs across Germany, using mailing lists, training events and postal mail. To ensure a diverse sample, we adopted a maximum variation sampling approach^11^, considering gender, region, and work experience (see Table 1). Moreover, we recruited GPs with personal experience of long-term sequelae after COVID-19 infection, adding first-hand experience to broaden our sample. Data collection continued until data saturation was reached, with a minimum of three participants per sampling criterion^12^.

**Table 1.** Summary of the maximum variation sample (n=31)

| Gender | Region of practice | Years practising as a GP | N |
| --- | --- | --- | --- |
| Women | urban | ≤10 | 4 |
|  |  | >10 | 4 |
|  | rural | ≤10 | 3 |
|  |  | >10 | 3 |
| Men | urban | ≤10 | 4 |
|  |  | >10 | 3 |
|  | rural | ≤10 | 3 |
|  |  | >10 | 3 |
| GPs experiencing long-term sequelae after COVID-19 infection |  |  | 4 |

### Data Collection

We followed a semi-structured interview guide to conduct the interviews. After conceptualising the interview questions based on the research questions and a systematic review of the literature, they were grouped into overarching topics. For each topic, an interview prompt and a set of follow-up questions were designed to elicit detailed responses^13,14^. The interview guide (see Supplement) covered the relevance of PCS in general practice, the role of GPs in patient care, diagnostic procedures, coding practices, sources of information, treatment strategies, interprofessional collaboration and unmet healthcare needs. We conducted a pilot test of the interview guide (n=2) and included the data in the final sample, as no significant changes were required.

JS (female, psychologist, PhD) performed the majority of the interviews, with LL (male, research assistant) and NP (female, psychologist, postdoctorate degree) each performing one interview. Both JS and NP were experienced in qualitative methods and LL, who was new to qualitative research, received extensive training and close supervision. Interviewers and participants were either unfamiliar or loosely acquainted, such as through professional associations. Prior to data collection, interviewers identified themselves as project staff and provided both verbal and written information about the study. There were no dropouts between recruitment and interview, and all participants gave written informed consent.

Participant characteristics were collected using a short sociodemographic questionnaire. We also assessed the number of patients with PCS, defined as the presence of sequelae three months or more following COVID-19 infection, seen by the GP. Field notes and audio recordings were taken during the interviews. We then produced verbatim transcriptions, which were quality controlled and anonymised.

### Data Analysis

MAXQDA software^15^ was used for data management and coding. We analysed the data using structuring qualitative content analysis according to Kuckartz^16^, which combines data-driven and concept-driven approaches. Based on the research questions, a deductive coding frame was developed that included code definitions, anchor examples and coding rules (see Supplement). Two coders (JS and LL) coded the first two transcripts together to establish a shared understanding of the coding frame and to refine coding rules. Subsequent coding was done independently, with discrepancies resolved through discussion with a third coder (ARA). Next, inductive subcategories were derived and the coding frame expanded accordingly, repeating the process and preparing thematic summaries. To ensure intersubjective comprehensibility, the interview guide, pilot test results and materials were presented and discussed in workshops with other researchers. In addition, participants were provided with the final report for feedback.

## Results

The interviews were conducted between August 2022 and July 2023 to account for changes in COVID-19 variants and health policies. The mean length of the interviews was 35 minutes. With two exceptions, the participants were based in the five federal states of northern Germany. Sample characteristics (*N*=31) are displayed in Table 2.

**Table 2.** Sample characteristics.

| Characteristics | $N=31$ |
| --- | --- |
| Age, mean (range) | 49.5 (33-74) |
| Years of experience as GP, mean (range) | 13.5 (1-45) |
| Gender, n (%) |  |
| Women | 16 (51.6) |
| Men | 15 (48.4) |
| Form of practice, n (%) |  |
| Individual practice | 6 (19.4) |
| Group practice (shared patients and expenses) | 4 (12.9) |
| Group practice (only shared expenses) | 16 (51.6) |
| Medical care centres | 3 (9.7) |
| Other | 2 (6.4) |
| Number of patients with a (confirmed or suspected) diagnosis of PCS, n (%) |  |
| $\leq 5$ | 6 (19.4) |
| 6-10 | 5 (16.1) |
| 11-15 | 11 (35.5) |
| 16-20 | 3 (9.7) |
| ≥21 | 6 (19.4) |

### Relevance of long-/post-COVID-19 syndrome in GP practices

GPs consider the perceived ‘quantitative’ importance of post-COVID to be low, with many noting a discrepancy between their experience and the high prevalence rates reported in some epidemiological studies. However, because of the severity of the cases, the issue is of greater ‘qualitative’ importance:

*“So it does play a role, but a very small one. For us, I’d say it’s more like a rare disease. So there are a few cases of post-COVID, but even more cases of long-COVID. Now, post-COVID, if we want to make that distinction, twelve weeks after infection, I have very few of them, but they are quite severely affected*.*” (GP_007)*

Ongoing symptomatic COVID-19 is a more common reason for seeking medical advice. Symptoms typically resolve spontaneously within a few weeks, a timeframe that is considered normal by GPs and therefore does not prompt them to take immediate action. However, patients often request a diagnostic evaluation because of concerns about possible long-term symptoms.

### Symptoms and consequences of illness

The main symptoms observed by GPs are reduced physical performance, fatigue and persistent cough. Other common symptoms include shortness of breath, chest tightness, palpitations, postural orthostatic tachycardia syndrome, fever, myalgia, joint pain and headache. Neurological symptoms have been reported, in particular ‘brain fog’ with difficulties in concentration, memory and word finding, as well as anosmia, ageusia and paraesthesia. Psychological effects such as loss of interest and depressed mood are also noted. These symptoms are generally characterised as ‘non-specific’. In some cases, PCS has been associated with the onset or exacerbation of other physical conditions such as myocarditis and diabetes.

GPs outlined a broad spectrum of functional impairments in patients with PCS, including reduced athletic performance, rapid physical and mental exhaustion and limited mobility. They recognise a high impact on social participation, such as reduced work capacity, prolonged absences, and job loss or changes. Reduced income, e.g., sickness or disability benefits, can be a source of financial hardship for patients. They also struggle with fulfilling social roles, including domestic responsibilities and maintaining personal relationships:

> „*What many patients have in common is that they are very limited in their ability to continue to work and participate in social life. (*…*) I feel that I can no longer take care of my family or anything like that. I’m no longer able to function as a mother or a father or at work*.*” (GP_005)*

### Perceived Patient Characteristics and Risk Factors

Opinions and experiences vary as to whether prevalence rates differ by sex. Most of the patients diagnosed with PCS initially had a mild acute infection but GPs also treat patients with long-term sequelae who had been hospitalised during the acute phase. Unvaccinated status is considered a risk factor for PCS. GPs report that the majority of patients are young or middle-aged:

*“It seems to me that younger patients are more likely to be affected and that older patients are much less likely to have these symptoms, especially after the first vaccination. Perhaps this is because they are no longer working and don’t have to deal with these kinds of demands*.*” (GP_025)*

There was overall agreement among participants that PCS is more common in patients with psychological distress or pre-existing mental health conditions:

> *“I always had the impression that they were more likely to be people who had been in treatment before or who had come to me for burnout, depression, depressive episodes or something like that. And when I did this search, it basically totally confirmed that*.*” (GP_031)*

### The GP’s role

GPs consider themselves the first point of contact for patients with persistent symptoms. Key tasks are to perform the initial diagnostic assessment, coordinate further diagnostic procedures, interpret specialist findings in the context of the patient’s history and symptoms and communicate these findings. They decide on specialist referrals and when to adopt a ‘watchful waiting’ approach. They also advise patients on self-management strategies while protecting them from non-evidence-based treatments. Managing functional limitations involves assessing their ability to work and facilitating rehabilitation as well as giving patients time through issuing sickness certificates and encouraging patience. They closely monitor patients’ health and symptoms. The GP’s role in caring for patients with PCS is characterised by uncertainty:

> *“I think that caring for post-COVID patients affects GPs in a special way. Because there is certainly a feeling of helplessness at times. We still don’t have many tools compared to, perhaps, knowing how to support depressed patients*.*” (GP_021)*

In the absence of immediate solutions, GPs emphasise the importance of taking patients’ concerns seriously, actively listening to them and providing reassurance.

### Diagnostic procedures, dilemmas and coding practice

When patients report persistent symptoms following COVID-19 infection, GPs conduct a comprehensive diagnostic assessment based on the principle of exclusion. This initial assessment includes a detailed medical history, physical examination, (resting or ambulatory) electrocardiogram (ECG), blood pressure monitoring and a battery of laboratory tests. GPs focus on identifying signs of inflammation and ruling out metabolic, cardiovascular and organ-specific conditions such as thyroid, liver and kidney disease. The aim is to recognise red flags for relevant differential diagnoses. If initial findings are normal – the typical scenario – GPs opt for a ‘watchful waiting’ strategy before considering further steps in order to avoid overdiagnosis.

After the initial assessment, further diagnostic tests are based on symptoms, with referrals to specialists depending on availability. These may include long-term and exercise ECGs, pulmonary function tests and imaging tests such as abdominal ultrasonography, chest computed tomography and echocardiography. Neurological examinations may be carried out to assess concentration and memory problems. Long waiting times for specialist assessment often lead to a lengthy diagnostic process. Psychological distress is usually assessed after the physical examination has been completed. By taking this sequential approach, GPs aim to increase patient receptivity to the possibility of psychological factors. Follow-up tests may also be conducted to provide additional reassurance.

A recurring theme was the difficulty in objectifying patients’ complaints with diagnostic tests. Diagnosis remains difficult due to the lack of clear markers. During or after the interview, many participants asked about the availability of PCS questionnaires. Distinguishing PCS from conditions with similar symptoms poses particular challenges, for example, in ascribing mental health problems either as sequelae of post-COVID or as pre-existing conditions:

> *“Then there is a huge field of, how do you say, rather non-specific symptoms: tired, exhausted, concentration problems, these things. And I find it very difficult to deal with them because I can’t really make a definitive attribution. So it’s impossible for me to decide whether it’s coincidental or causal*.*” (GP_018)*

The interviews revealed varying coding practices. The diagnosis of post-COVID-syndrome (ICD-10: U09.9) is usually made by GPs after a comprehensive diagnostic evaluation and the three-month time criterion. Sometimes, main symptoms (e.g. dyspnoea, fatigue) or exclusion diagnoses are coded. GPs tend to take a more conservative approach to coding, preferring to record it as a suspected diagnosis. On the one hand, fear of stigma and diagnostic uncertainty, particularly the possibility that there may be another unrecognised cause, as well as ambivalence towards the diagnosis, are reasons for GPs not to code. Some see coding as unnecessary or even counterproductive, as the true potential for chronicity is unclear and diagnostic labelling may hinder improvement. On the other hand, coding enables specific therapy and validates symptoms as a diagnosis, providing reassurance to patients:

“*The most important thing was to take the patients and their complaints seriously. And to code the illness. To say that they are not pretending, but (*…*) have an official diagnosis. You had to tell people that, too. It’s also coded here, it’s in the international code, and you’re not a simulant or hypochondriac*.*” (GP_028)*

### Interprofessional collaboration

Almost all participants reported collaborating with other health professionals to care for patients with PCS. Collaboration for diagnostic purposes involves specialists in cardiology, pulmonology, radiology, psychotherapy/psychiatry, neurology, otolaryngology, immunology and rheumatology. Collaboration for treatment purposes also includes physiotherapists, occupational therapists, rehabilitation sports groups and rehabilitation clinics.

Participants identified insufficient communication as a barrier to interprofessional collaboration. Diagnostic tests by specialists often return normal results with no further suggestions for treatment, leaving GPs with the sole responsibility for treatment decisions:

> *“So the reports from the specialists are usually quite rudimentary anyway, and then they just rule it out, okay, it’s nothing cardiovascular, period. Fine. But then I’d like the specialists to tell me what they do with people who have palpitations or what the pulmonologist does with people who have shortness of breath. But they just send them back to me and then I’m left here again with no idea what to do with them*.*” (GP_016)*

Some GPs reported being active in PCS networks or other local professional groups. Where formal or informal networks are in place, they facilitate communication about patients and swift referrals to colleagues with expertise in the specific clinical presentation. In most regions, however, such networks are lacking.

### Treatment strategies

#### Treatment in primary care

Rather than standardised regimens, treatment approaches in primary care vary according to symptomatology. GPs tend to offer more frequent and longer consultations. They explain the limited evidence available and the strong likelihood of spontaneous remission within 12 months. GPs stress the importance of the patient-provider relationship in achieving successful outcomes. They encourage patience and address feelings of hopelessness while helping patients cope with functional limitations:

> *“I feel like a lot of it is a mixture of encouragement and activation and support in the sense of psychological stabilisation and then exercising patience with them until things finally get better*.*” (GP_017)*

Self-management strategies include pacing, gradually increasing physical activity, avoiding overexertion, eating a healthy and nutritious diet and quitting smoking. There were reports of the benefits of olfactory training as a self-directed intervention. Pharmacological interventions mainly target physical symptoms such as cough and hypertension. Some GPs recommend dietary supplements such as vitamin C or B12. GPs also play a role in prevention, advising rest during the acute phase of infection. Participants considered the use of Paxlovid® to prevent PCS and discussed a possible link between booster vaccinations and improvement in symptoms, although this was solely based on single observations.

#### Secondary care

GPs recognise barriers to accessing mental health care, especially long waiting times and patients’ reluctance to seek help for fear of stigmatisation. GPs often take proactive steps to address this reluctance by advocating for a comprehensive understanding of illness that includes psychosocial factors and highlighting the relevance of coping mechanisms. Physical therapy is commonly prescribed, with GPs reporting that patients benefit from exercise, respiratory therapy, manual therapy and heat therapy. According to them, the aims should be to encourage patients to exercise in a safe and controlled way, to enable them to integrate exercise into their daily lives, and to rebuild confidence in their physical abilities. By comparison, occupational therapy receives less attention from GPs due to lower familiarity, with concentration problems emerging as the main indication in the interviews. GPs recommend rehabilitation exercise and support groups to promote social interaction and peer support. Overall, the availability of services is limited and expertise in PCS is often lacking.

#### Rehabilitation

In Germany, medical rehabilitation programmes require an application process and the approval and authorisation of the social insurance provider. GPs play a central role in initiating rehabilitation through proactive discussions and preparing the necessary documentation. However, determining the optimal time to apply for rehabilitation is challenging. Although the approval process is lengthy, suggesting an early initiation, GPs are cautious about starting too early, especially in cases of severe fatigue, to avoid overwhelming patients. Experience of rehabilitation outcomes varies widely, from significant improvement in symptoms to deterioration as a result of overexertion and early withdrawal from the programme. Transferring learned skills to daily life can be difficult, making long-term benefit uncertain. GPs facilitate patients’ return to work, but the timing is equally complex. They often question whether the indirect benefits of illness, in particular for those with recurrent sickness absence, might impede progress. From their perspective, returning to work is a gradual process, requiring adjustments such as reduced hours, pace and remote options, with no guarantee of success.

#### PCS clinics

GPs hold different opinions about referring patients to PCS clinics. While some advocate for increased collaboration with specialist facilities, others believe that usual care suffices. They point to difficulties in accessing such clinics and long waiting times, as well as concerns about a potential ‘diagnostic spiral’ leading to patient distress from incidental findings. This may also increase the focus on negative somatic sensations or create false expectations of treatment. Furthermore, lack of comprehensive patient history complicates symptom attribution and individualised treatment planning. Despite reservations, regular follow-up from specialists relieves pressure on GPs and validates patient concerns. Many feel able to manage without PCS-specialist care and reserve referrals for complex cases.

### Information resources

GPs rely on various sources of information on PCS. Most consult recent studies in scientific journals and attend training programmes. They frequently search the internet, use commercial medical information services and refer to the resources of professional organisations. Peer consultation, whether within formal structures or informally, is crucial for exchanging information. Although the German Guideline for Long/Post-COVID^17^ is widely used as a source of information, it is not universally embraced:

„*In principle, I like it. Because it presents things very clearly. And I also like the fact that it was adapted very quickly. (*…*) But it still doesn’t help me to sort out the criteria, so to speak, or to write the application for rehab. Or to help decide for the individual. Where should they go next? That’s what I miss. Well, I don’t think that the guideline can be that detailed. It has to be generic enough to be generally applicable*.*” (GP_006)*

Furthermore, there is a demand for improved information services for patients, potentially through an online portal, brochures, a central hotline or direct counselling. GPs expressed a desire for feedback and support in managing complex cases, potentially from PCS clinics.

## Discussion

### Summary

This is one of the first studies to explore GPs’ experiences of managing patients with PCS. While GPs perceive PCS prevalence as rather low, they often find themselves engaged in longer and more frequent consultations with patients affected by PCS. Fatigue and reduced physical performance are the main symptoms reported by GPs, and PCS is often seen in patients with a history of psychological distress. Patients experience a wide range of functional limitations and significant psychosocial consequences. GPs play a crucial role in diagnosis, usually performing the initial diagnostic assessment followed by a ‘wait and see’ approach and symptom-based testing. There is a tendency for a conservative approach to coding. GPs use various sources of information, including scientific journals and guidelines. They provide symptom-focused treatment, refer patients to other services where available, and place emphasis on the role of communication and the professional relationship. Questions arise about the utility of PCS clinics due to the lack of diagnostic markers and causal treatment. Interprofessional collaboration is crucial, yet challenged by communication barriers and the novelty and complexity of PCS diagnosis and treatment.

### Strengths and limitations

Although the study is specifically contextualised within primary care in Germany, we hypothesise that the findings on aspects such as symptoms as well as diagnostic and treatment strategies may have wider applicability and could potentially benefit other healthcare systems as well. However, there may have been a bias in the participation of GPs towards those with a greater interest in the subject matter. To mitigate these limitations, we sought to maximise the diversity of accounts by ensuring a heterogeneous distribution of participants in terms of gender, region, years in practice and first-hand experience, and we consider thematic saturation to have been reached as inductive category building was exhausted by the end of the analysis^12^.

### Comparison with the existing literature

The results align with previous findings, confirming the significant psychosocial burden of PCS on patients’ quality of life^18–21^. GP’s observations of symptoms are broadly consistent with the current literature^22–25^. Despite their high prevalence in epidemiological studies^26–28^, sleep disturbances were rarely mentioned in the interviews, indicating a potential gap for targeted interventions. Furthermore, gastrointestinal problems, more common in post-COVID patients than in the general population^29^, were not explicitly addressed.

The literature often suggests a positive association between PCS and older age, although potential confounders are debated, as older age is associated with increased prevalence of chronic comorbidities, risk of severe disease progression and mortality^30–32^. In a study by Bachmeier et al.^8^, German GPs reported that most patients were middle-aged or young, possibly reflecting the heightened visibility of functional limitations due to increased work and family responsibilities in these age groups.

The findings on the role of the GP are consistent with recent research highlighting their central role in PCS care^8,33^. This is reflected in the recently published Long COVID Policy of the German Federal Joint Committee^34^, which outlines the GP’s responsibility for structured initial assessment and coordination of further diagnostics and treatment. With a strong emphasis on communication in their approach to care, GPs prioritise what their patients are looking for: A sense of understanding and empathic support in coping with persistent symptoms and uncertainty^35^.

### Implications for research and clinical practice

At present, the diagnosis of PCS lacks clear operationalisation, covering a wide range of clinical pictures^36–38^. In order to standardise the diagnostic process, it seems necessary to define clear diagnostic criteria and severity categories. There is a need to develop practical assessment tools that include lesser known symptoms and to provide differential diagnostic protocols. Biomarker research is also of major interest. It should be noted, however, that patients may be less open to potentially helpful psychosomatic treatment if they are fixated on finding a somatic cause for their illness. This is not surprising, as patients may be concerned about the stigma associated with attributing their symptoms to psychological factors alone^39^. Training clinicians to communicate diagnostic uncertainty may therefore be helpful. At the same time, studies are needed to explore the relationship between PCS and psychological distress. Health policy measures are needed to foster interprofessional collaboration, e.g. by helping to establish or strengthen local networks. Access to psychotherapy, group treatments and specialised physical and occupational therapy needs to be improved. Attention should be given to the design of rehabilitation programmes to best support patients’ return to work^40^.

GPs advocate for better informational resources to be made available to patients. The German Ministry of Health’s Long COVID initiative has launched a website, but its uptake and acceptance by patients is yet to be determined. Additionally, GPs seek expert advice to manage complex cases, potentially from PCS clinics. Given the relatively small number of post-COVID patients in primary care, it is likely that their expertise will develop more slowly than in specialised settings^9^, supporting the practical value of this proposal.

Continued research in close collaboration with clinical practice and health policy is essential to further improve the care of patients with PCS^5,37^. This requires a coordinated effort from all stakeholders. Therefore, the results of this study will be discussed in implementation workshops with experts from different groups (patients, primary care, rehabilitation and payers) to inform recommendations for further action.

## Data Availability

Participants in this study did not give written consent for their data to be made public, so no supporting data is available due to the sensitive nature of the research. Coding examples can be found in the coding frame.

## Acknowledgments

The authors would like to thank David Janssen and Sarah Jess for their time transcribing interviews, as well as the participants who took part in the interviews.

## Ethical approval

Ethical approval for this study was obtained from the local ethics committee at the Center for Psychosocial Medicine of the University Medical Center Hamburg-Eppendorf (approval no. LPEK-0483).

