## Supplement for "A Qualitative Interview Study of General Practitioners’ Experiences of Managing Post-COVID-19 Syndrome"

### Supplement 1: Interview guide

#### Warm-up/perceived relevance/role

- Could you please tell me a little about yourself?
  - How long have you been working as a GP?
  - What is your speciality or focus?
- What is your motivation for participating in our study?
- What role has post-COVID played in your practice so far?
- How do you perceive your role as a GP when it comes to providing care for patients with post-COVID?

#### Symptoms/diagnosis/coding

- Could you share a case of a patient with post-COVID that you have encountered in your practice? Starting from the initial signs of the disease, could you describe the diagnostic process and the steps you took?
  - Please tell me about a patient you have treated whose situation was quite different.
- How do you code and categorise such symptoms?

#### Typical case or personal experience

- What is the typical profile of the patients with post-COVID syndrome you see in your practice?
  - What are the limitations they experience in terms of daily activities, participation, and overall quality of life?

  **OR**
- In the questionnaire, you mentioned that you personally experience lingering symptoms after COVID-19 infection. Could you share your own experience with these symptoms?

#### Treatment

- How do you approach the treatment of patients with post-COVID syndrome?
  - Which treatment strategies have you found to be effective in managing post-COVID symptoms?
  - Have you encountered any treatment strategies that have been less successful?
  - Do you offer any specialized services or interventions specifically tailored for patients with post-COVID syndrome? If so, could you provide more details about these services?

#### Collaboration

- Could you share your experience working with other healthcare professionals and facilities in providing care for your patients with post-COVID syndrome?

    - If not collaborating with other professionals: What changes would need to occur for you to refer patients to specialist services?
- What has been your experience with implementing rehabilitation measures for your patients with post-COVID syndrome?
- Are you part of a post-COVID network?
    - If so, could you tell me about your experience with it?
- What other types of care or services would you like to see made available for your patients with post-COVID syndrome?
- From your perspective, what gaps exist in the current provision of care for patients with post-COVID syndrome?

**Recognition of post-COVID syndrome as an occupational disease**

- Have you encountered cases where post-COVID syndrome was recognised as an occupational disease?
    - If so, what were the unique aspects or differences compared to other payers?

**Needs in the GP practices**

- Does treating patients with post-COVID syndrome increase your workload?
    - How do you account for these additional costs?
- When considering the case you described earlier, how did you gather information about the diagnosis and treatment?
    - How helpful have these resources been for you so far?
- Are you familiar with the clinical guideline on long/post-COVID syndrome?
    - How useful do you find it in your practice?
- Are there any specific areas where you have unanswered questions or would like more information?

**Conclusion**

- Is there anything else relevant to the outpatient care of post-COVID syndrome that we have not covered in our discussion?

Supplement to:

Schulze et al.: Managing Patients with Post-COVID Syndrome: A Qualitative Study of General Practitioners' Experiences

**<u>Supplement 2: Coding frame</u>**

**Code System**

| | |
|---|---|
| 1 (d) Perception of the GP's role | 74 |
| 2 (d) Relevance of long-/post-COVID syndrome in GP practices | 48 |
| 3 (d) Symptoms, risk factors and consequences of illness | |
| 3.1 (i) Perceived patient characteristics and risk factors | 40 |
| 3.2 (i) Symptoms | 100 |
| 3.3 (i) Consequences of illness | 73 |
| 4 (d) Diagnosis and coding | |
| 4.1 (i) Diagnostic procedures and principles | 74 |
| 4.2 (i) Dilemmas in diagnosis | 44 |
| 4.3 (i) Coding | 50 |
| 5 (d) Treatment strategies | |
| 5.1 (i) Non-pharmacological primary and secondary care | 79 |
| 5.2 (i) Pharmacological primary and secondary care | 25 |
| 5.3 (i) Prevention | 7 |
| 5.4 (i) Rehabilitation | 58 |
| 5.5 (i) Specialised care | 28 |
| 5.6 (i) Need for additional support/treatment services | 67 |
| 6 (d) Interprofessional collaboration | 81 |
| 7 (d) Information resources and training | |
| 7.1 (i) Used resources | 44 |
| 7.2 (i) Acceptance of the guideline | 32 |
| 7.3 (i) Information needs & gaps | 64 |

**NOTES** (d) = deductive categories, (i) = inductive categories

Supplement to:

Schulze et al.: Managing Patients with Post-COVID Syndrome: A Qualitative Study of General Practitioners' Experiences

**1 (d) Perception of the GP's role**

Code definition: All statements relating to the GP's own understanding of their role in the care of patients with long-/post-COVID syndrome are coded here. Statements referring to a form of medical 'helplessness' or 'inadequacy' in relation to the GP's own role should also be included.

Anchor example: "And I see myself as the first point of contact for coordinating diagnostic assessments that may have already been initiated or still need to be carried out. And I am often the person who issues the sick note. And then the reports to / So the sickness benefit ends. I have to send them to the medical service of the health insurance company. And then the future is sealed, so to speak. How this will continue with their ability to continue working, with their working capacity and so on." (001, Pos. 15)

Coding rules: Also related to the transitory period from acute COVID-19 infection to prolonged infection to post-COVID syndrome.

**2 (d) Relevance of long-/post-COVID syndrome in GP practices**

Code definition: This category includes all descriptions of the perceived significance (e.g. also frequency) of the condition (both based on confirmed diagnoses and suspected diagnoses by doctors and patients) in GP practice.

Anchor example: Interviewer: "Which role has this issue played in your practice so far?" - Interviewee: "It plays a minor role SO FAR in the practice because we have very few patients with post-COVID syndrome. You can really count them with one hand." (010, Pos. 5-6)

Coding rules: Not referring to acute COVID-19 infection.

**3 (d) Symptoms, risk factors and consequences of illness**

Code definition: Refers to information on symptoms, perceived patient characteristics and risk factors as well as limitations to patients' participation, activities and quality of life. It also describes when pre-existing conditions manifest as a result of a COVID-19 infection.

Anchor example: "Then it was really the case that they had coronavirus and it was a very severe coronavirus infection. They were bedridden for several weeks. And one patient, who is 27, really could not get out of bed. Then she developed depression because she was no longer able to work. And then it went on for half a year. In between, she also had really severe physical limitations. Tachycardia, so she also had a massive cardiovascular response." (010, Pos. 10)

Coding rules: Also descriptions of prolonged infections with progression to post-COVID syndrome.

**3.1 (i) Perceived Patient Characteristics and Risk Factors**

Code definition: This code describes perceived risk factors that are associated with an increased likelihood of developing long-term symptoms after a COVID-19 infection, e.g. certain pre-existing conditions or the way the infection is treated. It also includes statements about frequently occurring characteristics of affected patients, e.g. sex or age.

Anchor example: "We have a patient who has now been back in rehab, who was impaired prior to that, with pre-existing conditions such as COPD, coronary heart disease and I would say a psychological component. So more of a depressive mood." (017, Pos. 11)

Coding rules: Also related to the transitory period from prolonged infection to post-COVID syndrome.

**3.2 (i) Symptoms**

Code definition: Includes statements on the symptoms of post-COVID syndrome.

Anchor example: "So I think these are the typical symptoms: Fatigue, difficulty concentrating, not so much actual physical pain, very rarely I see people who report muscle pain, and also a prolonged period. But otherwise difficulty concentrating, tiredness, fatigue. What else can I think of? Sometimes a cough, although to be honest you have to say that EVERY cold causes or can cause a lingering cough." (027, Pos. 10)

Coding rules: Also related to the transitory period from prolonged infection to post-COVID syndrome.

**3.3 (i) Consequences of illness**

Code definition: Includes information on the negative consequences of post-COVID syndrome in terms of patients' participation, activities and quality of life.

Anchor example: "Climbing stairs was extremely difficult, getting out of bed, tiredness, feeling exhausted all the time, aching limbs and muscles for months. Then actually went on sick leave for several weeks, then tried to work, failed, then went on sick leave again for several months, then worked and unfortunately got coronavirus again. Not great either, but then things got worse again. So rather more intense long-COVID symptoms. She has since recovered. Recovered in the sense that she can work again, but she says she still has not been able to achieve her performance from before the first coronavirus infection and is quickly exhausted and tired and she just about manages her job and cannot really do much more. Where she used to be able to meet up with friends or do more chores and so on, it is very, very difficult for her now." (029, Pos. 10)

Coding rules: Also related to the consequences of illness during the transitory period from prolonged infection to post-COVID syndrome.

**4 (d) Diagnosis and coding**

Code definition: This category includes all descriptions of the diagnostic procedures and coding behavior as well as related challenges for suspected long-/post-COVID syndrome (both related to confirmation and exclusion) or diagnostic procedures that were not initiated with the suspicion of long-/post-COVID syndrome but resulted in this diagnosis. This code is also used for all descriptions of diagnostic procedures and codes used by other practitioners.

Anchor example: "And, of course, we checked her thoroughly. Including lung function, sent to a cardiologist, extensive laboratory diagnostics, myocarditis ruled out. The pulmonologist did not find anything except that she is practically less physically fit. So she has this exertional dyspnoea. She has a high resting heart rate, she gets out of breath quickly. She simply has significant physical limitations. But it is difficult to objectify this. She has no, she has no obstruction, no restriction. It is, yes. It is simply this long-COVID." (008, Pos. 12)

Coding rules: Refers not only to long-/post-COVID syndrome, but also to diagnosis of exclusion or differential diagnostics.

**4.1 (i) Diagnostic procedures and principles**

Code definition: This category includes all descriptions of the diagnostic procedure for suspected long-/post-COVID syndrome (both related to confirmation and exclusion) or diagnostic procedures that were not initiated with the suspicion of long-/post-COVID syndrome but resulted in this diagnosis. This code is also used for all descriptions of diagnostic procedures performed by other practitioners.

Anchor example: "So for up to four weeks, if they still experience symptoms, I assume that it is normal, in quotation marks, for viral infections and if they only have a cough, I try to treat it first with inhalation and so on, before I then go in the direction of examining lung function, checking the heart and all these things." (024, Pos. 44)

Coding rules: Refers not only to long-/post-COVID syndrome, but also to diagnosis of exclusion or differential diagnostics.

**4.2 (i) Dilemmas in diagnosis**

Code definition: This category includes all descriptions of the challenges related to diagnostic procedures for suspected long-/post-COVID syndrome (both related to confirmation and exclusion).

Anchor example: "Then there is a huge field of, how do you say, rather unspecific symptoms: tired, exhausted, concentration problems, these things and I find it very difficult to deal with them because I cannot really make a causal attribution. So it is impossible for me to decide whether it is coincidental or causal. And I think that makes it very, very difficult." (018, Pos. 10)

Coding rules: Refers not only to long-/post-COVID syndrome, but also to diagnosis of exclusion or differential diagnostics.

**4.3 (i) Coding**

Code definition: This category includes all information on which (ICD) codes are used for the present symptoms, including patients with suspected long/post-COVID syndrome. This also applies to information on the conditions and causes for which the diagnosis is assigned or not assigned.

Anchor example: "Then I always include the neurological symptoms. In this case, we coded it as dysaesthesia, I think, and also post-COVID. That's because we can't code post-COVID alone, but always together with the symptoms that come with it." (024, Pos. 36)

Coding rules: Refers not only to long-/post-COVID syndrome, but also to diagnoses of exclusion or differential diagnoses.

**5 (d) Treatment strategies**

Code definition: This code includes statements that address symptomatic treatment and treatment strategies (e.g. specific measures such as medication, pacing) – whether successful or unsuccessful. Supportive interventions and self-management are also included. In addition, statements relating to (the support of) interventions to restore the patient's ability to work and their occupational rehabilitation are taken into account. Statements on treatment in long-COVID outpatient clinics are also coded here. The prevention of long-/post-COVID and chronification is also included in this category.

Anchor example: "So I think there are actually only two patients who were really in rehab specifically for this and I would say that the one patient I mentioned at the beginning stopped twice. So I would say that her experience was rather bad. However, she then also/there were also studies that came to the conclusion that it was perhaps even too much that the patients were doing and that actually reinforced her belief that it was all too early with the rehab and too MUCH and that she should scale it back again. So I don't think she herself was convinced during rehab that it was the right thing for her. The other patient's experience was relatively good and I've now heard from another lady, who I don't treat in the practice, that she had a VERY good experience with rehab. So I would say that the spectrum is relatively broad, i.e. the experiences with rehab." (021, Pos. 18)

Coding rules: To be distinguished from diagnostics and collaboration (in particular communication between the professionals involved).

**5.1 (i) Non-pharmacological primary and secondary care**

Code definition: This code includes statements that address non-pharmacological treatment and management strategies for patients – successful or unsuccessful – in primary and secondary care. It also includes supportive interventions and self-management.

Anchor example: "I actually have the feeling that a lot of it is a mixture of encouragement and activation and support in the sense of psychological support and then exercising patience with them until things finally get better." (017, Pos. 37)

Coding rules: To be distinguished from diagnostics and collaboration (in particular communication between the professionals involved).

#### 5.2 (i) Pharmacological primary and secondary care

Code definition: This code includes statements that refer to the pharmacological treatment of long-/post-COVID symptoms.

Anchor example: "And then, of course, from the first symptoms, which affected the respiratory tract, we used the Budesonide spray or Spiriva inhaler. Spiriva inhaler, I didn't want any additional bronchial irritation from a dose inhaler with propellant gas, so they were all only given the inhalation powder and then we increased the dose accordingly and told the patient, so not just twice a day, but you can do up to four times a day and then two inhalations, because with these doses in the microgram range, not milligram but microgram range, nothing happens. And then they more or less administered the dosage themselves according to their symptoms and found it quite good that they could influence their symptoms a little themselves." (026, Pos. 24)

Coding rules: To be distinguished from prevention.

#### 5.3 (i) Prevention

Code definition: This includes all statements that refer to the prevention of post-COVID syndrome in people who are acutely ill or already have a prolonged course of the disease.

Anchor example: "And I, I don't know, maybe we actually avoided a bit of Long-COVID, because we also gave Paxlovid relatively frequently to patients at risk." (023, Pos. 112)

Coding rules: The information can refer to COVID-19 in general as well as to long-/post-COVID syndrome.

#### 5.4 (i) Rehabilitation

Code definition: This code includes statements related to (supporting and initiating) measures to restore patients' ability to participate and return to work. This includes rehabilitation, work integration programmes, social participation and support measures. It also covers classification as a disability. Also coded here are statements about home care services to compensate for the patient's impairments, even if they are covered by other payers.

Anchor example: "So I think there are actually only two patients who were really in rehab specifically for this and I would say that the one patient I mentioned at the beginning stopped twice. So I would say that her experience was rather bad. However, she then also/there were also studies that came to the conclusion that it was perhaps even too much that the patients were doing and that actually reinforced her belief that it was all too early with the rehab and too MUCH and that she should scale it back again. So I don't think she herself was convinced during rehab that it was the right thing for her. The other patient's experience was relatively good and I've now heard from another lady, who I don't treat in the practice, that she had a VERY good experience with rehab. So I would say that the spectrum is relatively broad, i.e. the experiences with rehab." (021, Pos. 18)

Coding rules: Questions and uncertainties related to socio-medical aspects are also coded here.

**5.5 (i) Specialised care**

Code definition: This code is used for statements relating to specialised long-/post-COVID care provided by facilities such as long-COVID outpatient clinics. It also includes care within the context of clinical studies.

Anchor example: "There are also long-COVID centres, but they are all at full capacity. Clinic X had to stop admitting patients for a while. I have no idea how they manage that. I imagine it's impossible at the moment because they're all very busy." (029, Pos. 8)

Coding rules: Special treatment programmes in secondary care such as respiratory therapy are not included. Lack of access to specialised care should be double-coded ('Need for additional support/treatment services').

**5.6 (i) Need for additional support/treatment services**

Code definition: This category includes information or statements that indicate that there is an identified need for additional treatment services. This includes, for example, descriptions of unmet need for medical care, specific therapies or treatments that patients need but do not receive, or where access is restricted. It also includes statements that indicate that there is no need for additional support.

Anchor example: "Basically, in many areas there is simply a lack of a rapid link to short-term psychotherapy. So that you can say: This has affected you. Let's quickly see if you can talk about it with someone other than me. But that's not so easy right now, is it?" (001, Pos. 24)

Coding rules: Only with regard to post-COVID syndrome. Double coding allowed in case of restricted access to treatments.

**6 (d) Interprofessional collaboration**

Code definition: This code includes the involvement, experiences and needs of GPs in (cross-sectoral) collaboration with other disciplines and/or professional groups, i.e. also the coordination efforts undertaken by GPs.

Anchor example: "And if I can't make any progress and the patient continues to feel unwell or is suffering, then I first try to get an appointment with a neurologist, which isn't easy either, but sometimes it does work out if you call and say, so what? Maybe you can pinpoint it a bit better in the personal phone call, then we can get appointments." (019)

Coding rules: The information can refer to long- or post-COVID syndrome.

**7 (d) Information resources and training**

Code definition: This category refers to all statements in which GPs describe the strategies they have used to obtain information or the extent to which they have read guidelines, specialist knowledge and scientific articles. This category also describes the need GPs see for research or the extent to which they participate in it. The need for additional information, further training or peer exchange also falls into this category.

Anchor example: Interviewer: "How do you get information about long or post-COVID?" - Respondent: "On the public pages. So I / In between, when I have the time and peace and quiet. So, of course, in discussions with colleagues, that's one thing. The other, of course, is through the websites, so I usually just type in a keyword or I get redirected and then through the various university clinics. I don't have a favorite / (laughs) Then 'AMBOSS' stays up to date. And you can see the references they use. And then I've looked at the studies. Along with the guidelines. Then, so to speak, exactly. What did we find on the internet / Oh, I've forgotten the name of the site. It's a site for general practitioners, actually a learning and information platform, 'Deximed'." (009)

Coding rules: The information can refer to COVID-19 in general as well as to long-/post-COVID syndrome.

**7.1 (i) Used resources**

Code definition: This category refers to all statements in which GPs describe which resources they have used so far to obtain information and further training on the topic of long-/post-COVID syndrome.

Anchor example: Interviewer: "How do you get information about long or post-COVID?" - Respondent: "On the public pages. So I / In between, when I have the time and peace and quiet. So, of course, in discussions with colleagues, that's one thing. The other, of course, is through the websites, so I usually just type in a keyword or I get redirected and then through the various university clinics. I don't have a favorite / (laughs) Then 'AMBOSS' stays up to date. And you can see the references they use. And then I've looked at the studies. Along with the guidelines. Then, so to speak, exactly. What did we find on the internet / Oh, I've forgotten the name of the site. It's a site for general practitioners, actually a learning and information platform, 'Deximed'." (009)

Coding rules: The information can refer to COVID-19 in general as well as to long-/post-COVID syndrome. At most, the guideline is mentioned in this category, but is covered in the code 'Acceptance of the guideline'.

**7.2 (i) Acceptance of the guideline**

Code definition: This category refers to all statements in which GPs report the extent to which they are familiar with the relevant guidelines, with a focus on the German S1 Guideline Long-/Post-COVID Syndrome. Any subjective assessments of the usefulness and comprehensiveness of the guideline are included here.

Anchor example: Interviewer: "And a guideline on long/post COVID has been developed. Do you know the guideline?" - Interviewee: "I think I scrolled through it once, but I cannot remember exactly. Well, I know it exists and I think I scrolled through it, but I am honestly not quite sure anymore." (016, Pos. 26-27)

Coding rules: The information can refer to COVID-19 in general as well as to long-/post-COVID syndrome.

**7.3 (i) Information needs and gaps**

Code definition: This category refers to all statements in which GPs describe the extent to which they see a need for additional information, further training or peer support for themselves or in general.

Anchor example: "So at the moment, one idea would be to have ONE contact person, for example for city A and city B in Germany or city C, it does not matter, and we have collected the information at an early stage, for example via the Association of Statutory Health Insurance Physicians or the Chamber of Physicians. So we get more technical information on how we can bill for something. All this nonsense with these test procedures alone. So who were we allowed to test, who was allowed to be on sick leave and for how long. The rules of the game were changed every five days. Crazy! Absolutely crazy, so there was money for 17,000 doctors to be informed about some nonsense on a daily basis, instead of taking the money and saying, well, we just need a center that offers us knowledge and know-how." (020, Pos. 56)

Coding rules: The information can refer to COVID-19 in general as well as to long-/post-COVID syndrome. Negative coding is also possible (no further information needed).
